# Parental perceptions of children’s physical activity participation: an exploration of satisfaction, school-based engagement, barriers to participation and preferred strategies for improvement

**DOI:** 10.64898/2026.05.30.26354500

**Authors:** Chidiebele Petronilla Ojukwu, Okolo Chukwunonyelum Judith, Adaeze I Onyekwelu, Chiedozie Eleje, Juliet Ekowa, Kadiree E Fatai

## Abstract

**Background:** Physical activity (PA) is essential for children’s physical, cognitive, and psychosocial development; however, many children do not meet recommended PA levels, particularly in low-and middle-income settings. Parents play a critical role in shaping children’s PA behaviours, yet limited empirical evidence exists regarding parental perceptions of PA participation, satisfaction, barriers, and improvement strategies within the Nigerian context.

**Methods:** A qualitative study was conducted in Enugu City, Nigeria, using in-depth semi-structured interviews with 20 parents of children enrolled in nursery, primary, and secondary schools. Participants were recruited purposively from community settings. Interviews were audio-recorded, transcribed verbatim, and analysed using reflexive thematic analysis following Braun and Clarke’s framework. Reporting adhered to the Consolidated Criteria for Reporting Qualitative Research (COREQ).

**Results:** Five themes were identified: (1) parents’ perceptions of children’s PA participation as context-dependent and variable; (2) satisfaction with PA opportunities existing along a continuum from high satisfaction to dissatisfaction; (3) perceived enablers, including accessible spaces, social support, parental involvement, and safety; (4) perceived barriers, notably time constraints, academic prioritisation, limited facilities, safety concerns, and parental availability; and (5) strategies for improvement, emphasising school-based reforms, parental engagement, community collaboration, and policy-level support.

**Conclusions:** Parental satisfaction with children’s PA opportunities was mixed and frequently conditional, with many participants expressing dissatisfaction related to academic prioritisation, limited time for physical education, and inadequate recreational facilities. The findings suggest that improving both satisfaction and participation may require strengthened school-based physical activity provision, greater parental engagement, and enhanced community infrastructure to support balanced child development.

## Introduction

Physical activity (PA) is widely recognised as a cornerstone of healthy child development, contributing significantly to cardiometabolic, musculoskeletal, cognitive, and psychosocial health [1–3]. Regular participation in physical activity supports the development of motor skills, improves emotional well-being, enhances social interaction, and contributes positively to academic functioning and quality of life [1,3–5]. Despite these well-established benefits, global evidence indicates that many children and adolescents do not achieve recommended levels of physical activity, with sedentary behaviours increasingly becoming common across different settings [6,7]. Insufficient physical activity among children has been associated with increased risks of obesity, type 2 diabetes, cardiovascular disease, poor mental health outcomes, and reduced overall well-being [2,8].

The World Health Organization (WHO) recommends that children and adolescents aged 5–17 years engage in at least 60 minutes of moderate-to-vigorous physical activity daily [2]. However, achieving these recommendations is influenced by multiple interconnected factors extending beyond children’s individual motivation or preferences. To conceptualise these influences, this study draws on the Socio-Ecological Model (SEM), which posits that health behaviours are shaped through interactions across individual, interpersonal, institutional, community, and policy-level environments [16,17]. Within the context of children’s physical activity participation, individual factors may include temperament, confidence, and health status; interpersonal influences may involve parental support, peer relationships, and family routines; institutional factors include school schedules, physical education provision, and extracurricular opportunities; while community and policy-level influences encompass neighbourhood safety, recreational infrastructure, and broader sociocultural priorities [16,17].

Among these interconnected influences, parents play a particularly important role in shaping children’s physical activity behaviours. Parents often act as “choice architects” by determining the value placed on active play, allocating time for participation, facilitating access to recreational opportunities, and investing resources in extracurricular activities [1,11]. Previous evidence suggests that children whose parents actively encourage or participate in physical activity with them are more likely to engage consistently in active behaviours [12]. Parental perceptions and attitudes therefore have important implications for children’s opportunities for participation and the extent to which physical activity is prioritised within daily family life.

In many low– and middle-income countries, including Nigeria, childhood physical activity participation is further influenced by broader institutional and environmental constraints. Emerging evidence suggests that school-based physical activity opportunities within Nigerian settings may be inconsistent and often constrained by competing academic demands [9]. In many urban educational environments, schools may place strong emphasis on academic performance, resulting in reduced opportunities for structured or unstructured physical activity. Long school hours, extensive homework schedules, and limited time allocated for physical education may contribute to increasingly sedentary routines among children [9,13].

Community-level conditions may further shape children’s participation in physical activity. Rapid urbanisation in many Nigerian cities has been associated with declining access to safe recreational spaces, increased traffic congestion, inadequate pedestrian infrastructure, and safety concerns that may restrict outdoor play opportunities [18,19]. These environmental conditions may limit parents’ willingness to allow children to engage freely in community-based physical activity, particularly within densely populated urban settings such as Enugu City. At the same time, socioeconomic and educational differences among families may influence parental expectations regarding school provision, extracurricular participation, and access to recreational resources [10,13].

Despite the recognised importance of parental influence on children’s physical activity participation, qualitative evidence exploring parental perceptions within the Nigerian context remains limited. Existing studies have focused predominantly on prevalence estimates or quantitative assessments of activity levels, with comparatively little attention given to how parents interpret, negotiate, and evaluate children’s physical activity opportunities across home, school, and community settings [9,13]. Understanding parental satisfaction, perceived barriers, and preferred strategies for improving participation is important for informing contextually relevant interventions and policies aimed at promoting active lifestyles among children [14,15].

This study therefore aimed to explore how parents in Enugu, Nigeria, perceive and interpret their children’s physical activity participation, including their satisfaction with existing opportunities, perceived barriers to participation, and preferred strategies for improvement. By examining these experiences within the broader socio-ecological context, the study seeks to contribute evidence that may inform family-centred, school-based, and community-level approaches to promoting physical activity among children.

## Materials and Methods

### Study Design

This study utilised a qualitative descriptive research design to explore parental perceptions of children’s physical activity participation across home, school, and community environments. A qualitative approach was considered appropriate because it allows for an in-depth and contextually grounded exploration of perceptions, experiences, and meanings that cannot be fully captured through quantitative methods [20,21]. The study was conceptually informed by the Socio-Ecological Model (SEM), which recognises that health behaviours are shaped through interactions across individual, interpersonal, institutional, community, and policy-level influences [16,17]. Reporting of the study was guided by the Consolidated Criteria for Reporting Qualitative Research (COREQ) checklist [22].

### Context and Setting

The study was conducted in Enugu City, the administrative capital of Enugu State in southeastern Nigeria. Enugu is a rapidly urbanising metropolitan area characterised by a mixture of public institutions, commercial centres, and residential communities. The educational environment within the city is often characterised by strong emphasis on academic achievement, with many schools implementing extended school hours, supplementary lessons, and substantial academic workloads. At the same time, urbanisation and infrastructural limitations may contribute to reduced access to safe recreational spaces and organised opportunities for physical activity among children. These contextual conditions made Enugu City an appropriate setting for exploring parental perceptions of children’s physical activity participation.

### Ethical Considerations

Ethical approval for the study was obtained from the University of Nigeria Teaching Hospital Health Research Ethics Committee (NHREC/05/01/2008B–FWA00002458–IRB00002323). Written informed consent was obtained from all participants prior to data collection. Participation was voluntary, confidentiality was maintained throughout the study, and participants were informed of their right to withdraw from the study at any stage without consequence.

### Participant Selection and Sampling Strategy

Participants were recruited using purposive sampling to identify information-rich individuals capable of providing detailed insight into the research question [23]. Eligible participants were parents or primary caregivers of children enrolled in nursery, primary, or secondary schools within Enugu City who were familiar with their children’s physical activity participation in school or community settings. Parents whose children had severe physical, cognitive, or psychological impairments that could substantially limit participation in physical activity were excluded to maintain focus on typical participation experiences within mainstream school contexts.

To ensure variation in perspectives, recruitment was conducted across diverse public settings commonly accessed by parents, including churches, marketplaces, shopping centres, and recreational areas within Enugu City. During recruitment, efforts were made to include parents from different educational backgrounds, occupational categories, school types (public and private), and children’s educational levels (nursery, primary, and secondary school). Recruitment continued until thematic sufficiency was achieved, whereby later interviews contributed additional depth and clarification rather than substantially new conceptual insights [24,25]. A total of 20 parents participated in the study.

### Data Collection Procedures

Data were collected using semi-structured in-depth interviews conducted between August and October 2025. The interview guide was developed by the research team based on the study objectives, relevant literature, and the socio-ecological framework underpinning the study. Questions explored parental perceptions of children’s physical activity participation, satisfaction with available opportunities, perceived barriers, and strategies for improving participation across home, school, and community settings. The interview guide was pilot-tested with two parents who were not included in the final study sample, resulting in minor modifications to improve clarity and flow.

To accommodate participants’ preferences and schedules, both telephone and face-to-face interviews were utilised. Fourteen interviews were conducted via telephone, while six interviews were conducted face-to-face at locations selected by participants to ensure privacy and comfort. All interviews were conducted in English and lasted between 30 and 45 minutes.

Data collection was conducted by a three-member research team comprising CO, KF, and AO, all of whom have training and experience in qualitative research. Prior to data collection, the team reviewed the interview guide collectively and discussed interviewing approaches to ensure consistency in questioning and probing techniques across interviews. CO served as the primary interviewer, while KF and AO assisted with field notes, interview coordination, and follow-up probing where necessary. Written and verbal informed consent was obtained from all participants prior to participation, and with participants’ permission, all interviews were audio-recorded to facilitate accurate transcription.

### Researcher Positionality and Reflexivity

The research team maintained ongoing reflexive engagement throughout the study to acknowledge how researchers’ backgrounds and assumptions may have influenced data collection and interpretation. The researchers are Nigerian physiotherapists and public health researchers with professional interests in child health, rehabilitation, and physical activity promotion. These backgrounds may have shaped assumptions regarding the importance of physical activity and concerns about increasingly sedentary lifestyles among children.

The researchers recognised that their professional identities could position them as authority figures during interviews, potentially influencing participants’ responses. To minimise this influence, interviews were conducted using open-ended, non-judgmental questioning, and participants were reassured that there were no right or wrong responses. Throughout data analysis, reflexive discussions and peer debriefing sessions were conducted to critically examine interpretations and ensure that findings remained grounded in participants’ accounts rather than researchers’ preconceptions.

### Data Analysis

Data were analysed using Reflexive Thematic Analysis (RTA) following the six-phase framework proposed by Braun and Clarke [24,25]. This approach recognises coding and theme development as interpretive and shaped through researchers’ active engagement with the data.

Audio-recorded interviews were transcribed verbatim, and all identifying information was removed to maintain confidentiality. The transcripts were read and re-read by the research team to facilitate familiarisation with the dataset. Initial codes were then generated inductively through close engagement with participants’ narratives. Coding focused on both explicit meanings and broader patterns related to parental perceptions of children’s physical activity participation.

The researchers subsequently engaged in collaborative reflexive discussions to explore patterns across the dataset and develop candidate themes. Rather than seeking statistical inter-coder agreement, these discussions were used to deepen interpretation, refine conceptual understanding, and strengthen analytic rigour. Themes were reviewed iteratively against the coded extracts and the full dataset to ensure coherence and meaningful representation of participants’ experiences.

The final themes were defined and refined through continued interpretive discussion within the research team. Illustrative participant quotations were selected to support the analytic narrative and demonstrate the relationship between the data and the study findings. Regular reflexive discussions and peer debriefing sessions were conducted throughout the analytic process to enhance credibility and ensure that interpretations remained grounded in participants’ accounts.

## Results

### Participant Characteristics

A total of 20 parents participated in the semi-structured interviews. The demographic and socioeconomic characteristics of the participants and their children are presented in Table 1. The sample represented a range of educational and family contexts within Enugu City.

**Table 1.** Socio-demographic characteristics of parents and children (N = 20) Five major themes were identified from the qualitative analysis: (1) parents’ perceptions of children’s physical activity participation, (2) parents’ satisfaction with physical activity opportunities, (3) perceived enablers of physical activity participation, (4) perceived barriers to physical activity participation, and (5) perceived strategies for improving children’s physical activity participation.

| Variable | Category | Frequency (n) | Percentage (%) |
| --- | --- | --- | --- |
| Parent's Age (years) | 31–40 | 7 | 35.0 |
|  | 41–50 | 8 | 40.0 |
|  | 51–60 | 5 | 25.0 |
| Parent's Gender | Male | 6 | 30.0 |
|  | Female | 14 | 70.0 |
| Educational Attainment | Secondary Education | 2 | 10.0 |
|  | Graduate (BSc/HND) | 8 | 40.0 |
|  | Postgraduate (MSc/PhD) | 10 | 50.0 |
| Occupational Category | Government Public Sector | 15 | 75.0 |
|  | Self-employed/Business Owner | 5 | 25.0 |
| School Type | Public/Government School | 7 | 35.0 |
|  | Private School | 13 | 65.0 |
| Child's Educational Level | Nursery School | 6 | 30.0 |
|  | Primary School | 4 | 20.0 |
|  | Secondary School | 10 | 50.0 |
| Child's Age (years) | 1–5 | 6 | 30.0 |
|  | 6–10 | 4 | 20.0 |
|  | 11–15 | 7 | 35.0 |
|  | 16–17 | 3 | 15.0 |

Regarding the educational level of children, half of the participants reported that their children were enrolled in secondary schools (n = 10, 50.0%), while 30.0% (n = 6) had children in nursery schools and 20.0% (n = 4) had children in primary schools. Reflecting this distribution, children’s ages ranged from early childhood to adolescence, with 35.0% (n = 7) aged 11–15 years, 30.0% (n = 6) aged 1–5 years, 20.0% (n = 4) aged 6–10 years, and 15.0% (n = 3) aged 16–17 years.

Most parents were aged between 41 and 50 years (n = 8, 40.0%), followed by those aged 31–40 years (n = 7, 35.0%) and 51–60 years (n = 5, 25.0%). The majority of participants were female (n = 14, 70.0%). Half of the participants held postgraduate qualifications (n = 10, 50.0%), while 40.0% (n = 8) were university graduates. Most participants were employed within the government public sector (n = 15, 75.0%), and the majority had children attending private schools (n = 13, 65.0%).

### Theme 1: Parents’ Perceptions of Children’s Physical Activity Participation

Parents described children’s physical activity participation as dynamic and highly dependent on context. Many participants reported that children behaved differently across home, school, and community environments, with some children appearing reserved within structured school settings but highly active at home.

“In school… he’s very quiet… but at home, he’s super active.” (P1, Female)

Several participants also explained that family routines and parental involvement strongly influenced participation in physical activity. Some parents described intentionally creating opportunities for active play and exercise within the home environment.

“At home and in church, the environment is structured; he participates because I intentionally involve him in exercises and activities with his siblings.” (P20, Female)

Children’s personalities were further perceived as shaping their engagement in physical activity. Parents frequently described children as energetic, shy, outgoing, or reserved, and interpreted these characteristics as influencing willingness to participate in active play or organised activities.

“I will describe her as super active… very active in both school and community.” (P14, Female)

Several parents further expressed concern regarding the frequency and consistency of participation in physical activity. Participation was often described as irregular and constrained by school schedules, fatigue, and competing academic demands.

“They leave home very early and return late, so they are usually too tired to play.” (P18, Female)

Parents’ accounts suggest that children’s physical activity participation is shaped by interacting influences across home, school, and community environments. Rather than being determined solely by individual motivation, participation appeared to vary according to family routines, school structures, and contextual opportunities available to children within their daily environments.

### Theme 2: Parents’ Satisfaction with Physical Activity Opportunities

Parents’ satisfaction with children’s physical activity opportunities varied considerably and existed along a continuum ranging from high satisfaction to dissatisfaction. To further illustrate this variation, participants’ responses were grouped into broad satisfaction categories (Table 2).

**Table 2.**
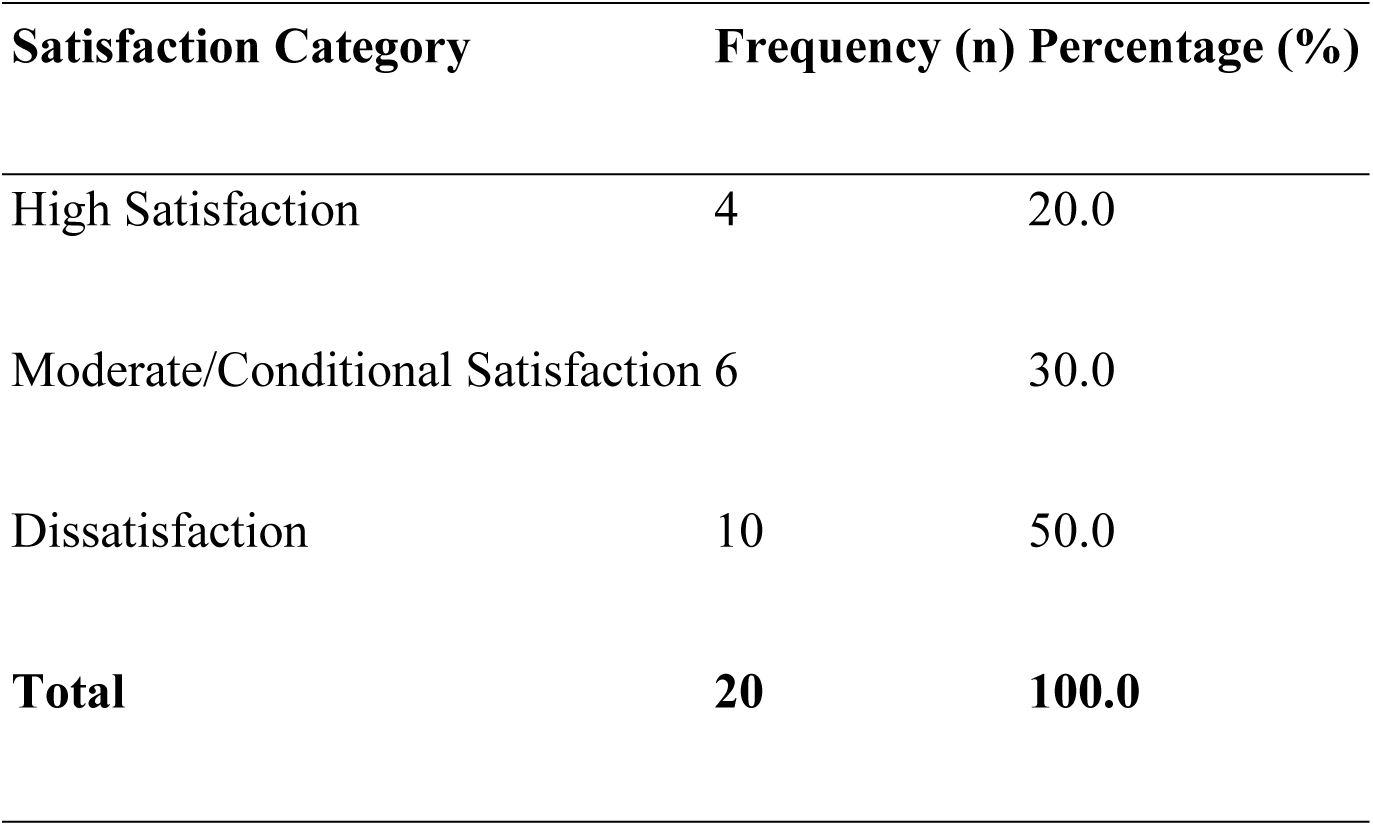
Parental satisfaction with children’s physical activity opportunities (N = 20) A small number of participants reported high satisfaction with school and community physical activity opportunities, particularly where schools provided organised sports programmes and structured physical education.

“I am very satisfied with the opportunities provided in school and community; I would rate it 10 out of 10.” (P17, Male)

Several participants expressed moderate or conditional satisfaction. Although they acknowledged the availability of some opportunities, they felt that the frequency and quality of activities remained limited.

“The school tries, but I wish they had more regular sports programmes.” (P5, Female)

“The activities are fair, but there is still not enough time for exercise because of academic work.” (P9, Female)

Many participants expressed dissatisfaction with current physical activity opportunities. Dissatisfaction was commonly linked to inadequate facilities, limited play spaces, safety concerns, and the perceived prioritisation of academics over physical development.

“I am not really satisfied because there is hardly enough time for play and physical activities.” (P8, Female)

“There are not enough safe places for children to play around here.” (P6, Male)

Across different satisfaction levels, parents consistently emphasised the importance of physical activity for children’s overall health, development, and well-being.

Parents’ satisfaction with physical activity opportunities appeared to reflect broader differences in school provision, environmental resources, and available support systems. Participants who reported higher satisfaction often described structured school programmes and supportive environments, whereas dissatisfaction was commonly associated with limited opportunities, inadequate facilities, and competing academic demands.

### Theme 3: Perceived Enablers of Physical Activity Participation

Parents highlighted multiple family, school, and environmental factors that supported children’s physical activity participation. At the family level, parental encouragement and active involvement emerged as important facilitators. Participants who valued physical activity often created opportunities for active play, encouraged outdoor activities, and attempted to reduce sedentary behaviours within the home.

“Parents are key… registering kids in activities and encouraging them to be active.” (P7, Female)

“I encourage my children to play outside whenever there is free time instead of sitting with phones.” (P12, Male)

School-related factors also emerged as important enablers. Parents whose children attended schools with structured sports activities, physical education periods, and basic recreational facilities perceived these opportunities as supportive of regular participation.

“The school has sports activities every week, and that helps because the children have time set aside for exercise.” (P11, Male)

Supportive social relationships were also perceived as facilitating participation. Peers, siblings, teachers, and extended family members were frequently described as encouraging children to participate in physical activities. Accessible and safe spaces for play further influenced participation. Parents reported that children were more likely to engage in physical activity where playgrounds, open spaces, or safe compounds were available.

“When children have space to run around and friends to play with, they are naturally more active.” (P15, Female)

The findings suggest that children’s physical activity participation is facilitated when supportive family practices, school structures, and safe environments operate together. Parents often felt more confident encouraging participation when schools provided structured opportunities and when children had access to safe spaces for active play.

### Theme 4: Perceived Barriers to Physical Activity Participation

Participants identified multiple barriers that limited children’s participation in physical activity across school, family, and community environments. Academic demands emerged as one of the most commonly reported barriers. Parents described long school hours, extra lessons, homework, and examination pressures as reducing the time and energy available for physical activity.

“By the time they return from school and extra lessons, they are already tired and still have homework to do.” (P18, Female)

“Most of their time is spent on school activities, assignments, and preparing for exams.” (P10, Female)

Several parents also perceived that schools prioritised academic achievement over physical education and recreational activities.

“The timetable is tight, and physical education is not regular, so it affects participation.” (P5, Female)

Community-level barriers were also frequently discussed. Parents expressed concern regarding inadequate recreational facilities, lack of playgrounds, traffic hazards, and neighbourhood safety concerns.

“There are no safe places for children to play around here anymore.” (P9, Female)

“Because of traffic and safety concerns, I prefer keeping them indoors most of the time.” (P13, Male)

Parental work schedules and household responsibilities were further described as limiting opportunities for supervision and active engagement with children. Some participants also identified child-related factors such as illness, shyness, screen time, and preference for indoor activities as barriers to participation.

“Sometimes he just prefers staying indoors with television and games.” (P3, Female)

The barriers described by parents reflected interconnected challenges across institutional, environmental, family, and individual levels. Academic pressures, limited recreational infrastructure, and safety concerns were frequently perceived as factors beyond parental influence, while family schedules and screen-related behaviours further constrained opportunities for regular physical activity participation. Parents often described difficulties balancing children’s academic responsibilities with opportunities for physical activity participation.

### Theme 5: Perceived Strategies for Improving Physical Activity Participation

Parents proposed several strategies for improving children’s physical activity participation across school, family, and community settings. School-based recommendations included increasing the frequency of physical education, improving sports facilities, organising regular sporting activities, and creating more balanced school schedules.

“Schools should create more time for sports and reduce the pressure from constant academic work.” (P6, Female)

“There should be more organised sports and better playground facilities in schools.” (P16, Male)

At the family level, participants highlighted the importance of parental involvement, role modelling, and reducing excessive screen time.

“Parents also need to participate and encourage children instead of allowing them to stay indoors with phones and television.” (P4, Male)

Community and policy-level recommendations included improving neighbourhood safety, creating parks and recreational centres, and supporting community-based physical activity programmes for children.

“Government should provide more safe recreational centres where children can play freely.” (P19, Female)

Parents’ recommendations highlighted the need for coordinated approaches involving families, schools, communities, and broader policy support. Participants generally perceived that improving children’s physical activity participation would require both behavioural support within families and structural improvements within school and community environments.

## Discussion

This study provided a contextually grounded qualitative exploration of parental perceptions regarding children’s physical activity participation in Enugu City, Nigeria. Guided by the Socio-Ecological Model (SEM), the findings demonstrate that children’s physical activity participation is shaped by interacting influences across individual, family, school, community, and broader environmental levels. Rather than being determined solely by children’s motivation or parental preference, participation was influenced by institutional academic demands, parental practices, environmental safety, and the availability of supportive recreational opportunities.

### The Role of Family and Parental Influence

The findings of this study highlight the important role parents play in shaping children’s physical activity participation. Parents described themselves as key facilitators of physical activity through encouragement, role modelling, structured family routines, and regulation of sedentary behaviours such as excessive screen time. These findings align with previous studies demonstrating that parental support, co-participation, and behavioural modelling are positively associated with higher levels of physical activity among children and adolescents [10–12].

Parents in the present study also perceived children’s physical activity behaviours as highly dependent on context. Children were described as behaving differently across home, school, and community settings, suggesting that participation is influenced not only by individual characteristics but also by the environments within which children interact daily. This finding supports socio-ecological perspectives which propose that health behaviours emerge through interactions between individuals and their surrounding social and physical environments [16,17].

The study further revealed that parents who intentionally created opportunities for active play within the home environment were more likely to report positive perceptions of children’s participation. However, parents frequently acknowledged that their efforts alone were insufficient when broader institutional and environmental barriers limited opportunities for participation.

### Academic Demands and Institutional Influences

Academic demands emerged as one of the most significant barriers to children’s physical activity participation. Parents consistently described long school hours, extra lessons, homework, and examination pressures as reducing the time and energy available for physical activity. These findings reflect concerns previously reported in studies suggesting that educational systems may increasingly prioritise academic achievement at the expense of physical activity opportunities [9,13].

Participants also perceived that some schools placed limited emphasis on physical education and recreational activities. Although a few parents expressed satisfaction with schools that provided organised sports programmes and structured physical education, many participants described school schedules as heavily dominated by academic activities. This finding suggests that institutional priorities may strongly influence opportunities for movement and active play among children.

The findings are particularly important within the context of urban Nigeria, where academic achievement is often viewed as an important pathway for social and economic advancement.

Parents frequently described difficulties balancing children’s academic responsibilities with opportunities for regular physical activity participation. This tension appeared to shape family routines, after-school schedules, and parental decisions regarding children’s daily activities.

### Environmental and Community-Level Barriers

Community and environmental conditions also emerged as important influences on children’s physical activity participation. Parents frequently expressed concerns regarding inadequate recreational spaces, traffic hazards, neighbourhood safety, and the declining availability of open play areas within urban communities. These findings are consistent with previous studies linking neighbourhood safety and access to recreational infrastructure with physical activity participation among children and adolescents [18,19].

Rapid urbanisation may contribute to reduced opportunities for outdoor play, particularly in densely populated urban environments such as Enugu City. Parents in the present study often perceived community environments as insufficiently supportive of active lifestyles, particularly where playgrounds, parks, or safe open spaces were unavailable. Safety concerns further influenced parental decisions, with some participants reporting that they preferred keeping children indoors to minimise exposure to traffic or neighbourhood risks.

These findings demonstrate how environmental conditions may indirectly shape family-level behaviours and parenting practices. Although many parents valued physical activity and attempted to encourage active behaviours, their efforts were often constrained by broader environmental conditions perceived to be beyond their control.

### Enablers and Opportunities for Participation

Despite the barriers identified, parents also highlighted several factors that supported physical activity participation. Structured school sports programmes, physical education classes, parental encouragement, peer relationships, and access to safe play environments were all perceived as important facilitators. These findings suggest that children’s physical activity participation may improve when supportive family practices, school structures, and environmental opportunities operate together.

Importantly, parents who reported higher satisfaction with physical activity opportunities often described schools with organised sports activities and accessible recreational facilities. This finding reinforces the importance of institutional support in promoting active lifestyles among children. It also suggests that schools may serve as important intervention settings for improving physical activity participation in urban Nigerian contexts.

### Implications for Practice and Policy

The findings of this study highlight the need for multi-level strategies aimed at improving children’s physical activity participation within urban Nigerian settings. At the institutional level, schools may benefit from policies that strengthen physical education programmes, increase opportunities for organised sports, and create more balanced schedules that accommodate both academic and physical development.

At the community level, improving access to safe recreational spaces, parks, and child-friendly infrastructure may help create environments that support active play and movement. Urban planning policies that prioritise pedestrian safety and recreational infrastructure may further encourage participation in physical activity.

Family-centred approaches may also be important. Public health interventions should support parents with practical strategies for encouraging active lifestyles within home environments, particularly in contexts where outdoor opportunities are limited. Encouraging family-based physical activities and reducing excessive screen time may contribute positively to children’s overall movement behaviours.

Overall, the findings suggest that improving children’s physical activity participation requires coordinated efforts involving families, schools, communities, and policymakers rather than relying solely on individual behaviour change.

### Strengths and Limitations

This study provides important qualitative insight into parental perceptions of children’s physical activity participation within an urban Nigerian context. The use of semi-structured interviews allowed participants to describe their experiences and concerns in depth, while the application of the Socio-Ecological Model provided a useful framework for understanding the interaction between individual, family, institutional, and environmental influences.

However, several limitations should be acknowledged. The participant sample was predominantly composed of highly educated parents employed within the government public sector, and many participants had children attending private schools. As such, the findings may reflect primarily urban middle-class experiences and may not fully represent the perspectives of lower socioeconomic or rural populations. Additionally, the study relied on parental self-report and did not include direct measures of children’s physical activity participation.

## Conclusion

This study demonstrates that children’s physical activity participation in Enugu City, Nigeria, is shaped by interconnected influences across family, school, community, and environmental contexts. Although parents generally recognised the importance of physical activity for children’s health and development, participation was frequently constrained by academic pressures, limited recreational opportunities, safety concerns, and environmental barriers.

The findings highlight the importance of adopting multi-level approaches to promoting physical activity among children. Strengthening school-based physical education, improving access to safe recreational environments, and supporting parents with practical strategies for encouraging active lifestyles may help improve physical activity participation among children within urban Nigerian settings.

## Data Availability

De-identified qualitative data supporting the findings of this study are available from the corresponding author upon reasonable request, subject to ethical and confidentiality considerations.

## Abbreviations

COREQ: Consolidated Criteria for Reporting Qualitative Research
PA: Physical Activity
PE: Physical Education
PHE: Physical and Health Education
PTA: Parent–Teacher Association
SES: Socioeconomic Status
UNEC: University of Nigeria, Enugu Campus
WHO: World Health Organization

## Acknowledgements

The authors would like to thank all parents who participated in the study for sharing their time and experiences.

## Authors’ contributions

CO conceived the original idea for the study. CO, OJ, OA, and EC contributed to the development and refinement of the research questions and study design. CO, OJ, and OA conducted the data collection. CO, KF, and AO participated in the data analysis and interpretation. KF drafted the initial manuscript. All authors critically reviewed the manuscript and approved the final version for publication.

## Ethics approval and consent to participate

Ethical approval for the study was obtained from the University of Nigeria Teaching Hospital Health Research Ethics Committee (NHREC/05/01/2008B–FWA00002458–IRB00002323). Participant recruitment and data collection were conducted between August 2025 and October 2025 in Enugu City, Nigeria. Written informed consent was obtained from all participants prior to participation in the study. All participants were adults and participated voluntarily. Participants were informed of their right to withdraw from the study at any stage without consequence. Confidentiality and anonymity were maintained throughout the study, and no monetary or material incentives were provided.

## Consent for publication

Not applicable.

